# Lung transplantation for pulmonary fibrosis secondary to severe COVID-19

**DOI:** 10.1101/2020.10.26.20218636

**Authors:** Ankit Bharat, Melissa Querrey, Nikolay S. Markov, Samuel Kim, Chitaru Kurihara, Rafael Garza-Castillon, Adwaiy Manerikar, Ali Shilatifard, Rade Tomic, Yuliya Politanska, Hiam Abdala-Valencia, Anjana V. Yeldandi, Jon W. Lomasney, Alexander V. Misharin, G.R. Scott Budinger

## Abstract

Lung transplantation can potentially be a life-saving treatment for patients with non-resolving COVID-19 acute respiratory distress syndrome. Concerns limiting transplant include recurrence of SARS-CoV-2 infection in the allograft, technical challenges imposed by viral-mediated injury to the native lung, and potential risk for allograft infection by pathogens associated with ventilator-induced pneumonia in the native lung. Additionally, the native lung might recover, resulting in long-term outcomes preferable to transplant. Here, we report the results of the first two successful lung transplantation procedures in patients with non-resolving COVID-19 associated acute respiratory distress syndrome in the United States. We performed smFISH to detect both positive and negative strands of SARS-CoV-2 RNA in the explanted lung tissue, extracellular matrix imaging using SHIELD tissue clearance, and single cell RNA-Seq on explant and warm post-mortem lung biopsies from patients who died from severe COVID-19 pneumonia. Lungs from patients with prolonged COVID-19 were free of virus but pathology showed extensive evidence of injury and fibrosis which resembled end-stage pulmonary fibrosis. Single cell RNA-Seq of the explanted native lungs from transplant and paired warm post-mortem autopsies showed similarities between late SARS-CoV-2 acute respiratory distress syndrome and irreversible end-stage pulmonary fibrosis requiring lung transplantation. There was no recurrence of SARS-CoV-2 or pathogens associated with pre-transplant ventilator associated pneumonias following transplantation in either patient. Our findings suggest that some patients with severe COVID-19 develop fibrotic lung disease for which lung transplantation is the only option for survival.

**Single sentence summary:** Some patients with severe COVID-19 develop end-stage pulmonary fibrosis for which lung transplantation may be the only treatment.

## INTRODUCTION

As of August 17, 2020, over 5.6 million people have been diagnosed with coronavirus disease (COVID-19) in the United States and over 173,000 have died (https://coronavirus.jhu.edu) with greater than 2.4 million active cases. In a significant proportion of patients, COVID-19 infection can progress to severe respiratory failure and the Acute Respiratory Distress Syndrome (ARDS) requiring mechanical ventilation^1,2^. The reported mortality of COVID-19 patients requiring mechanical ventilation is between 20-40% despite optimized supportive care^3,4^.

Lung transplantation is a life-saving treatment for a variety of end-stage lung diseases^5^. Annually, over 2700 transplants are performed in the United States with a 1-year survival over 90% and 3-year survival over 75% (www.srtr.org). Several concerns limit the use of lung transplantation as a therapy for patients with severe ARDS secondary to COVID-19. First, there is a concern that SARS-CoV-2 or superinfecting pathogens associated with viral pneumonia in the native lung might recur in the allograft. Second, severe vascular and pleural damage secondary to SARS-CoV-2 infection might create technical barriers to transplant, increase ischemic time and worsen outcomes. Third, the severe deconditioning associated with prolonged mechanical ventilation, sedation and neuromuscular blockade might complicate recovery after transplant. Fourth, and perhaps most importantly, there is uncertainty as to whether the lung can repair after severe SARS-CoV-2 pneumonia, resulting in long-term outcomes better than lung transplantation. We successfully performed lung transplantation in two patients with SARS-CoV-2 pneumonia requiring prolonged mechanical ventilation and extracorporeal membrane oxygenation (ECMO) support in whom recovery was determined to be highly unlikely. We examined the explanted native lungs and the lungs from patients who died from COVID-19 using histology, smFISH for SARS-CoV-2 RNA, tissue clearing to image extracellular matrix organization, and single cell RNA-Sequencing. Lung histology and extracellular matrix imaging revealed evidence of severe fibrosis in the explanted lungs. SARS-CoV-2 viral transcripts were not detected in the explants using smFISH and there was no evidence of recurrent SARS-CoV-2 infection in the allograft. Striking similarities were observed when single cell RNA-Seq data from lung tissue from explants and those who expired from prolonged COVID-19-induced respiratory failure were mapped onto published RNA-Seq datasets generated from the lungs of patients undergoing transplantation for pulmonary fibrosis. We found that lung disease after severe and prolonged SARS-CoV-2 acute respiratory distress syndrome shared pathologic and molecular features with pulmonary fibrosis requiring transplantation, suggesting lung transplantation may be the only option for survival in these patients.

## RESULTS

### Case Descriptions

#### Case 1

A latinx female in her 20s presented with two weeks of poor appetite, gastrointestinal discomfort, fevers, cough, dyspnea on exertion, and pleuritic chest pain and was diagnosed with COVID-19. Upon presentation she was severely hypoxemic, refractory to oxygen therapy, and was emergently intubated. Nasopharyngeal swab and bronchoalveolar lavage were positive by PCR for SARS-CoV-2. She received mechanical ventilation according to ARDS net guidelines^6^ using a higher PEEP/lower FiO2 strategy. Her PaO2/FiO2 ratio was <150 repeatedly so she was ventilated prone for a total of three 16 hour periods in the first week of her illness^7^. Bronchoscopic sampling of the alveolar space was performed after intubation and when clinical co-infection or super-infection was suspected. The results of quantitative cultures and multiplex PCR detection for respiratory pathogens (BioFire FilmArray Respiratory 2 Panel) was used to manage antibiotics over the course of her hospitalization. Despite these interventions, the patient’s oxygenation worsened and she was placed on veno-venous extracorporeal membrane oxygenation (ECMO). Her course was complicated by a right sided pneumothorax requiring multiple pleural tubes and the development of *Serratia marcescens* pneumonia with left lower lung necrosis (Figure 1A,B). Later in her course she developed elevated pulmonary arterial pressures (71/49 mmHg) and echocardiography showed moderate right ventricular dysfunction with severe tricuspid regurgitation and congestive hepatopathy. Systemic anticoagulation was initiated but was complicated by a hemothorax and a liver capsular bleed necessitating emergent exploratory laparotomy. Throughout her medical course, she received broad-spectrum and pathogen-directed antibiotics, remdesivir, hydroxychloroquine, tocilizumab, and convalescent plasma. Her lung compliance, gas exchange, chest radiographs, and chest tomogram showed worsening or no improvement over the course of her ICU stay. Beginning 6 weeks from the onset of mechanical ventilation, bronchoalveolar lavage fluid PCR tests for SARS-CoV-2 were repeatedly negative. ECMO wean was attempted over the course of several weeks with no success and she was listed for lung transplantation. She received bilateral lung transplantation using central veno-arterial ECMO. Severe dense vascular adhesions were noted bilaterally with severe distortion of hilar planes and reactive lymphadenopathy (Figure 1C). Following transplantation, she received maintenance immunosuppression using tacrolimus, mycophenolate mofetil, and prednisone. In the ensuing two weeks, she was separated from VV-ECMO as well as mechanical ventilation and was discharged for inpatient rehabilitation at 4 weeks from transplantation. Her neurocognitive status and muscular strength improved rapidly following the transplantation. Two months after transplantation, the patient is home with oxygen saturations above 98% on room air and reports independence in activities of daily living.

**Figure 1.**
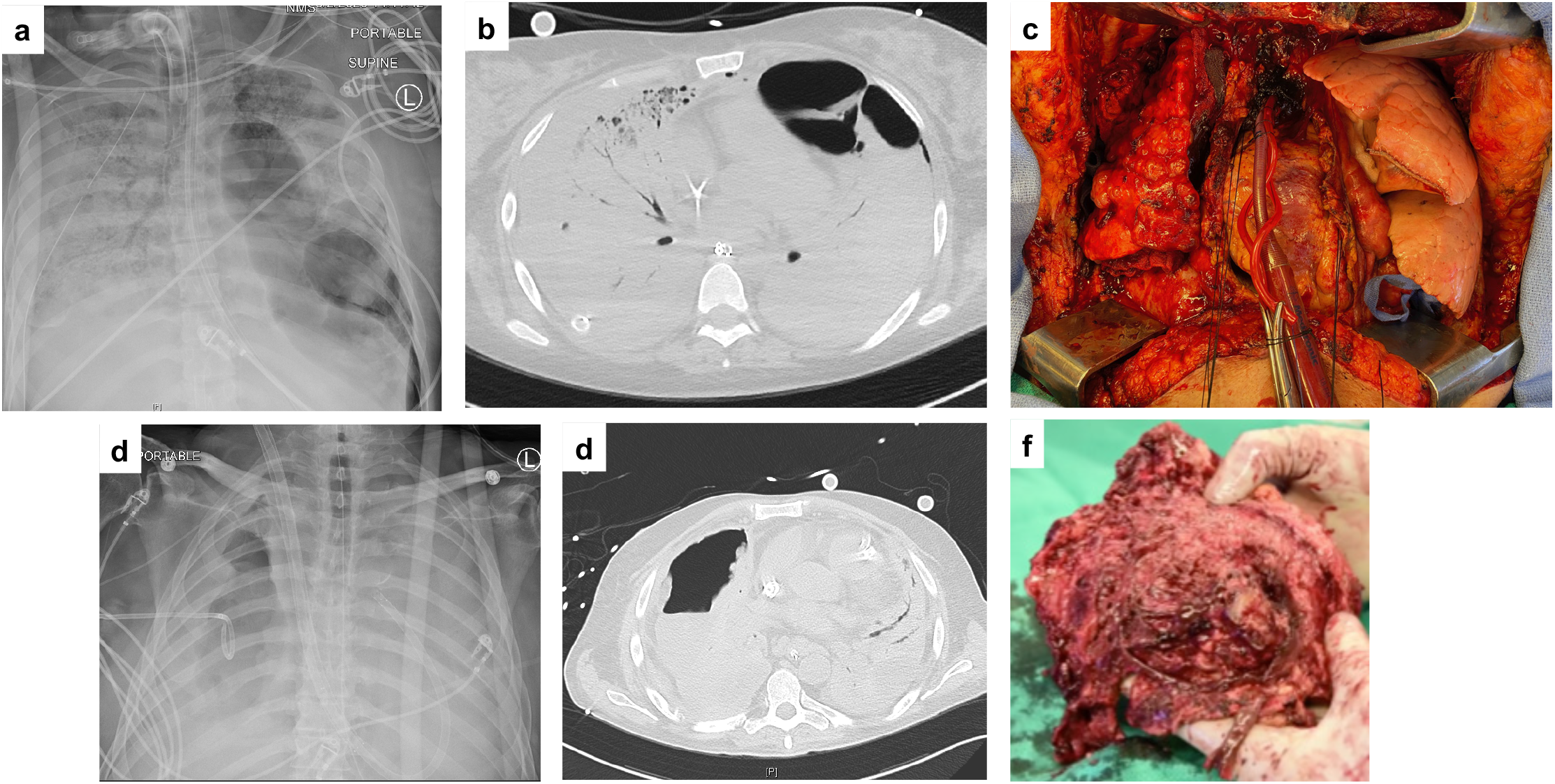
Radiographic and intraoperative findings in COVID-19 transplant recipients. **a**. Pre-transplant chest radiograph (Day 38 of onset of ARDS) of Lung Transplant Case 1 reveals opacification of the right lung and a left lower lobe necrotic cavity attributed to *Serratia marcascens* pneumonia. A tube thoracostomy was required to treat right spontaneous hemothorax and peumothoraces. **b**. Severe ARDS and lower lobe necrosis was confirmed on cross-sectional computed tomography. **c**. Intraoperative image obtained after revealing contrasting features between the two. The native lung reveals extensive cystic changes. The pericardium is open to provide access to the ascending aorta for cannula placement. **d**. Pre-transplant chest radiograph (Day 98 of onset of ARDS) of Lung Transplant Case 2 revealing bilateral lung opacifications and a necrotic cavity on the right lung attributed to *Pseudomonas aeruginosa* pneumonia.. A chest tube is in place to treat bronchopleural fistula. **e**. Computed tomography demonstrating severe ARDS and development of necrotic cavity in right lung. **f**. Freshly explanted right lung with extensive pleural inflammation and loss of identifiable anatomic planes.

### Case 2

A man in his 60s was placed on VV-ECMO for severe COVID-19 induced ARDS before transfer to our institution. During his course, he received remdesivir, convalescent plasma, and antibiotics. His course was complicated by recurrent *Pseudomona aeruginosa* pneumonia, hemothorax and empyema requiring thoracotomy and lung decortication (Figure 1D,E). His lung compliance and oxygenation failed to improve and he could not be weaned from VV-ECMO. Beginning 4 weeks after hospitalization, repeated bronchoscopic sampling of BAL fluid was negative for SARS-CoV-2 by PCR. After 100 days on VV-ECMO support, there were no signs of lung recovery and he was listed for lung transplantation. The intraoperative findings were similar to the first patient but increased complexity was encountered due to liquefactive necrosis from secondary *Pseudomonas* pneumonia and the prior lung decortication procedure associated with complete loss of normal mediastinal tissue planes (Figure 1F). At 30 days following transplantation, the patient is breathing on room air with oxygen saturations over 97%. He continues to improve with regards to neurocognitive status and muscular strength at an inpatient rehabilitation facility.

### Histologic Evaluation of Lung Tissue

The lungs from both patients were edematous and heavy (595.7/476.3 and 496.2/548.2 g right/left lung, Cases 1 and 2). In Case 1, the external surface of the lung explant had a nodular surface and dense fibrous adhesions between the lobes. In both patients, there were dense pleural adhesions, and the first patient had visible pleural blebs and cysts (Figure 1A). In both of the explants, large cavities with necrosis were present and post-operative cultures confirmed the presence of the bacterial pathogens in these cavities that were detected on BAL PCR and culture preoperatively.

Lung tissue from both patients and those obtained during warm autopsies included several shared features that are consistent with other autopsy series from patients with COVID-19^8^. There were areas of diffuse alveolar hemorrhage (Figure 2B and Supplemental Figure 1) as well as acute bronchopneumonia from secondary bacterial infection. In other areas the lungs showed various stages of a pattern consistent with an acute interstitial pneumonitis including acute neutrophilic infiltrates within the interstitium and alveolar spaces, interstitial expansion by fibrosis, bronchiolization of alveoli and areas of microscopic honeycomb changes. (Figure 2C-I). Rare microthrombi were observed in both lungs, some with recanalization (Figure 2J). Case 2 developed infarction and necrosis secondary to larger thrombi. Alveolar macrophages within the airspaces stained positive for iron, confirming the presence of alveolar hemorrhage (Figure 2K, L). Sections from the lung explant of Case 1 showed multiple cystic structures in various stages of formation. Early formation consisted of acute bronchilolitis with intra luminal neutrophils (Figure 2M), others were lined by histiocytic cells with chronic inflammation and giant cells (Figure 2N) while more mature cysts were devoid of inflammatory cells and some were associated with complete fibrosis (Figure 2O). These cysts were distinct histologically from the cysts of honeycomb lung. While the cystic changes we observed might reflect changes associated with prolonged mechanical ventilation, similar structures were observed in another patient who died from COVID-19 after deciding to forgo mechanical ventilation. Some of these cystic changes with histiocytic and giant cell reaction are reminiscent of pneumatoceles possibly from peripheral airway destruction as a result of viral or bacterial infection.

**Figure 2.**
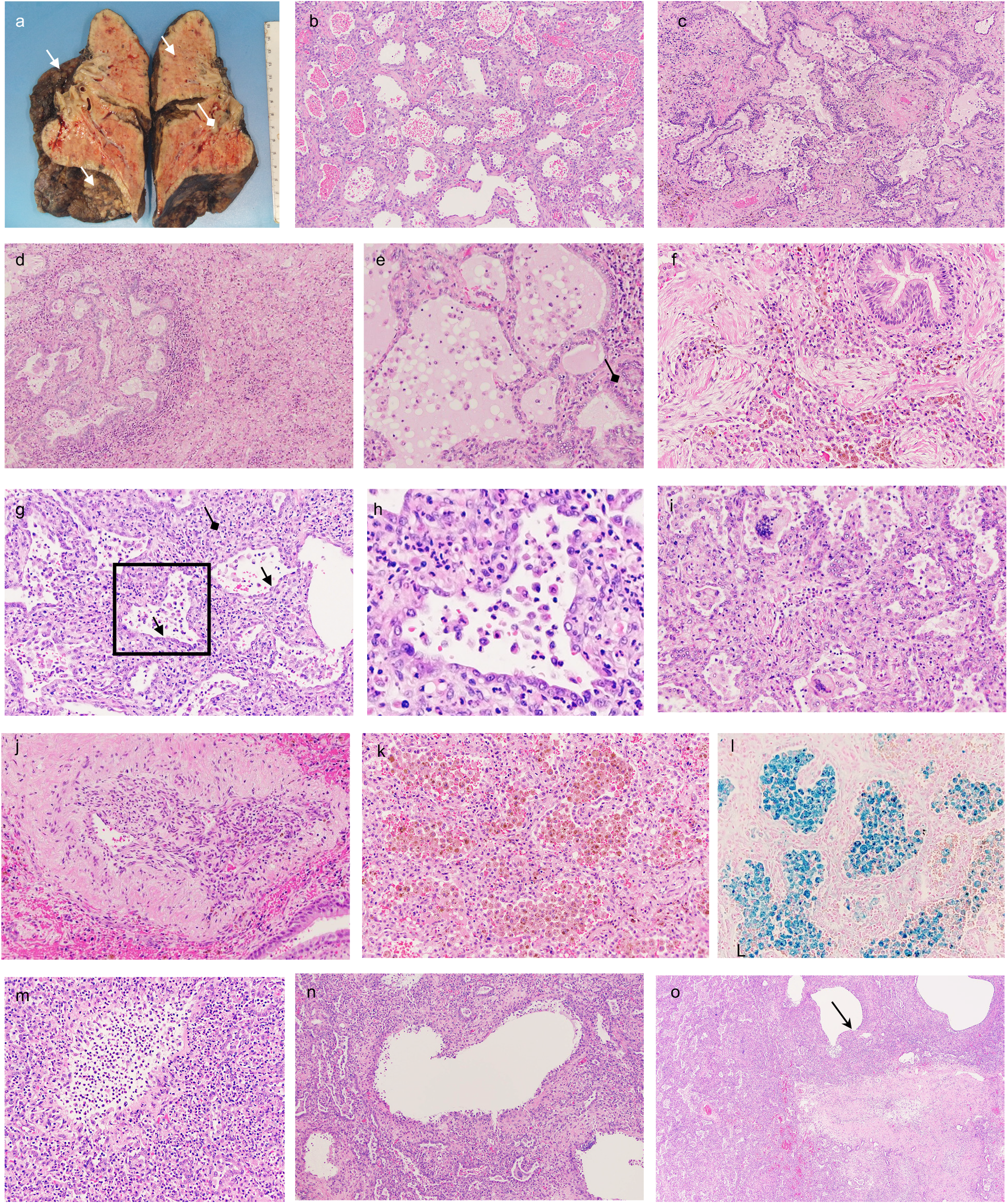
Common histologic features of the lung explants. **a**. Section of the lung at autopsy shows numerous cystic changes, fibrosis, pleural reaction and airway hemorrhage. **b**. Bronchiolitis at 200x. **c**. 100X. Interstitial fibrosis, septal expansion by inflammatory cells and myofibroblasts. **d**. 100X Honeycomb changes. **e**. Neutrophilic abscess formation. Dense fibrosis. Bronchiolitis. **f**. A 100x. Cystic airspaces with hemorrhage. Acute and chronic interstitial pneumonia. **g**. 200x. DAD with Type II pneumocyte hyperplasia. Multinucleate giant cells. Alveolar hemorrhage. Organizing pneumonia with dense fibrosis. **h**. 100x. Organizing DAD/pneumonia. Dense collagen whorls with numerous fibroblasts. Chronic inflammation. **i**. 100x. Organizing DAD/pneumonia. Obliteration of the normal architecture. Airspaces are filled with Type II pneumocytes, macrophages, fibrin membranes, red blood cells and fibroblasts. Dense collagen is present.

### Imaging of explanted and warm post-mortem lung tissue

The explants were obtained 40 and 100 days after the initiation of mechanical ventilation. Prior to transplant, BAL fluid was tested repeatedly for the presence of SARS-CoV-2 by a polymerase chain reaction test and were negative. During replication, the positive RNA strand of the SARS-CoV-2 virus is reverse transcribed to a negative strand, which serves as a template for hundreds of positive strands. The presence of both positive and negative strands in a cell is suggestive of ongoing viral replication. Accordingly, we performed smFISH using a commercial system RNAscope with probes designed to detect positive and negative RNA strands encoding SARS-CoV-2. While a control sample from lung tissue obtained from the patient who had forgone therapy for respiratory failure secondary to COVID-19 showed many positive and fewer negative strands of SARS-CoV-2 RNA, we were unable to detect virus using this technology in the explants from the two lung transplants (Figure 3A-C). Post-operative PCR performed on bronchoalveolar lavage confirms lack of SARS-CoV-2 recurrence.

**Figure 3.**
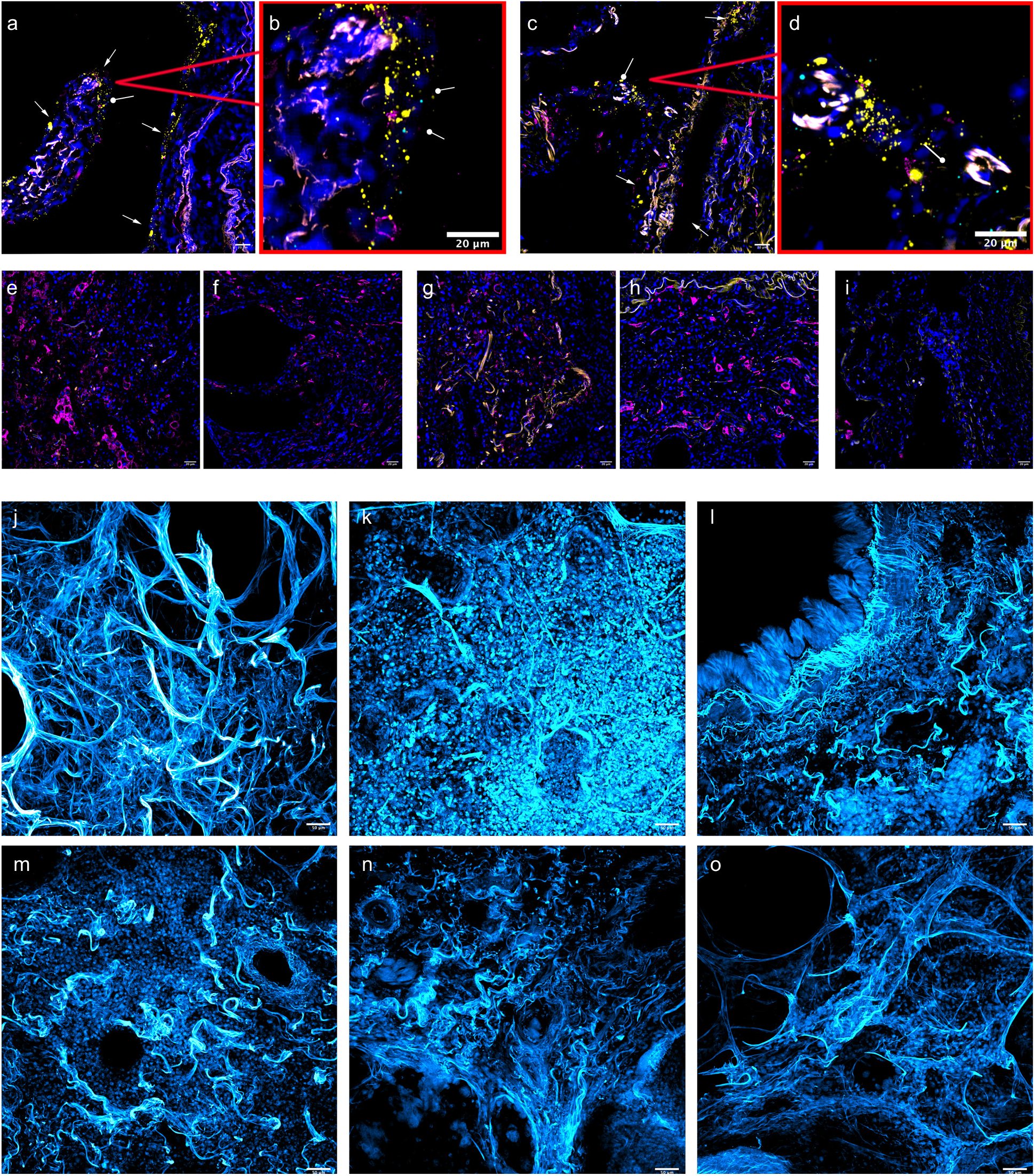
Single molecule (sm)-FISH for replicating SARS-CoV-2 and three-dimensional imaging of matrix organization in cleared human lung sections from patients with severe COVID-19. For **a-d**, the following RNAscope/IHC stains were used: nuclear (blue, network trained), SARS-CoV-2 sense mRNA (yellow), SARS-CoV-2 antisense mRNA (Cyan), CD206 (magenta). **a,b**. RNAscope of autopsy tissue from a patient who declined interventions for COVID-19 induced respiratory failure. Positive viral RNA (yellow) was detected in the alveolar epithelium (arrow), negative sense RNA was also detected (circular arrow) **c,d**. Another region from the same patient. **e,f**. RNAscope of the explanted lung from Lung Transplant Case 1 shows yellow autofluorescence but no staining for SARS-Cov-2 positive or negative strand RNA in left (left) or right (right) lung. Rare RNAscope background is seen. **g,h**. RNAscope findings from the explanted lung from Lung transplant Case 2, similar to Case 1. **i**.. RNAscope of post-mortem lung biopsy (patient 1). For **j-o**, hoescht-33342 was used for nuclear identification (Cyan Hot). In all images, matrix autofluorescence is shown at 20x magnification. **j**. Healthy lung tissue control. **k**. Explanted lung from lung transplant case 1 (COVID-19). **l**. Post-mortem biopsy from patient 1 (COVID-19) **m**. Post-mortem biopsy from patient 2 (COVID-19. **n,o**. Peripheral lung biopsy samples from lung explants obtained from patients with idiopathic pulmonary fibrosis.

Pulmonary fibrosis is characterized by the development aberrant and disorganized collagen deposition in the alveolar regions of the lung. We sought to compare the three-dimensional matrix organization of end stage lungs from patients with SARS-CoV-2 pneumonia with patients undergoing lung transplantation for idiopathic pulmonary fibrosis. We adapted protocols for tissue clearing (SHIELD) described in the brain to generate and image matrix organization in 3-D. A biopsy from the healthy donor lung tissue (Figure 3E, Supplemental Video 1) shows bands of collagen surrounding terminal airways with wispy lines defining the airways and airspaces, which are devoid of cells or matrix. Images from the explanted lung from Case 1 show a complete absence of matrix organization with punctate islands of cells surrounding fibrotic airway regions (Figure 3F). A similar pattern was observed in biopsies from two patients who died from severe COVID-19 pneumonia after a similar duration of mechanical ventilation whose families consented to a post-mortem lung biopsy for research (Post-mortem cases presented in the Supplemental Materials) (Figure 3G-H). This disorganized pattern of matrix organization was also observed in lung sections from the explants of two patients with idiopathic pulmonary fibrosis undergoing lung transplantation (Figure 3I-J). Three dimensional images can be observed in the supplemental movie (Supplemental Movie 1).

### Transcriptional Analysis of explant lungs and warm autopsies

Single cell RNA-Sequencing (RNA-Seq) has been applied to the lungs of patients with pulmonary fibrosis using tissues obtained at the time of lung transplantation to create transcriptional atlases of fibrosis^9-11^. These analyses have identified several cell populations that are seen in the fibrotic lung but are absent in normal lungs, many of which have been causally implicated in the development of pulmonary fibrosis in animal models. We mapped single cell RNA-Seq data obtained from the first transplant patient and both the post mortem biopsy samples along with two additional samples from normal donor lungs onto these cell atlases. In total, we sequenced 189,560 cells from which we identified all of the cell populations in previously published cell atlases of the normal lung and pulmonary fibrosis (Supplemental Figure 3)^9^. Consistent with our smFISH results, we did not detect SARS-CoV-2 positive or negative transcripts in our dataset (not shown). The dataset can be explored using interactive web browser at https://www.nupulmonary.org/covid-19/.

An emerging literature suggests that pulmonary fibrosis begins with disordered alveolar epithelial repair. Specifically as alveolar type 2 cells differentiate into alveolar type 1 cells, a transitional cell population of aberrant basaloid cells characterized by the expression of *TP63, KRT5, KRT17, LAMB3, LAMC2, VIM, CHD2, FN1, COL1A1, TNC, HMGA2* and several senescence markers (*CDKN1A, CDKN2A, CCND1, CCND2, MDM2, SERPINE1*) accumulate during fibrosis where they may contribute to disease pathogenesis through their expression of *TGFB1, ITGAV* and *ITGB6*^12,13^. A population of these cells with similar features was observed in all four patients who died or required transplant for COVID-19-induced respiratory failure but was absent from control lungs (Figure 4a-c).

**Figure 4:**
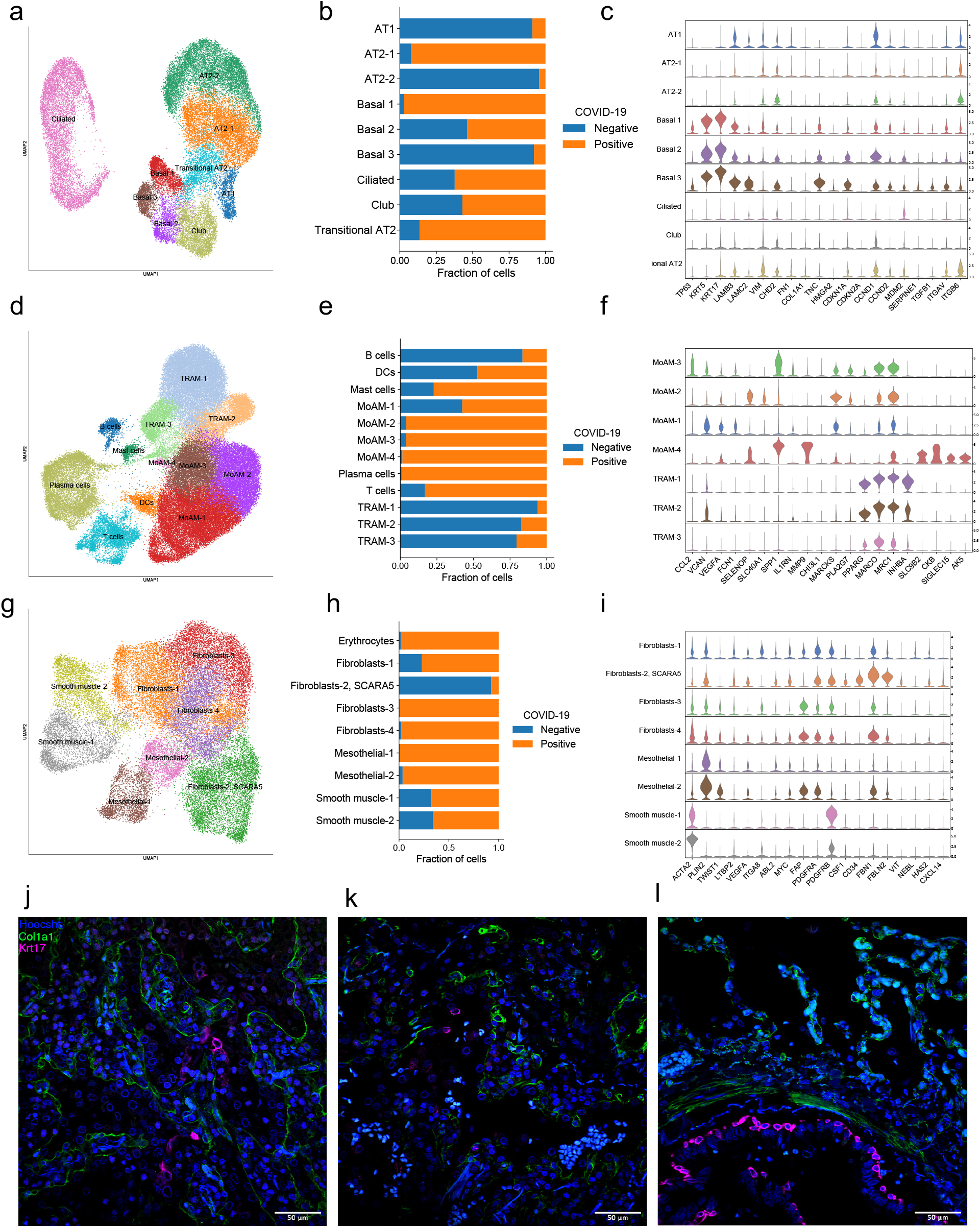
Single cell RNA-seq identifies similarities between end-stage pulmonary fibrosis and organizing pneumonia resulting from COVID-19. **a,d,g**. UMAP plots showing individual populations of epithelial (**a**), immune (**d**), and stromal (**g**) cells. **b,e,h**. Stacked bar plots illustrating cluster composition by the COVID-19 diagnosis for each of the lineages: epithelial (**b**), immune (**e**), and stromal (**h**). **c,f,i**. Violin plots illustrating expression of the selected marker genes characterizing individual subclusters: epithelial (**c**), immune (**f**), and stromal (**i**) cells. **j,k,l**. Immunofluorescent images from (**j**) Lung transplant case 1, (**k**) Lung transplant case 2 and (**l**) normal lung. KRT17 cells (magenta) are found in areas of fibrosis near COL1A1 positive fibroblasts (green). Nuclei are blue (Hoechst). Scale bar indicates 50 µm.

Disordered epithelial repair likely results in the continuous recruitment of monocyte-derived alveolar macrophages to the alveolar space that are necessary for the development of fibrosis in animal models^14^. Indeed, we observed abundant macrophages in the airways and alveolar spaces of our explants (Figure S2). In humans profibrotic monocyte-derived alveolar macrophages are characterized by expression of *SPP1, ILRN, MMP9, CHI3L1, MARKS*, and *PLA2G7* as well as reduced expression *PPARG, MARCO* and *MRC1*, which mark tissue-resident homeostatic alveolar macrophages present in the normal lung^9,12,13,15^. Macrophages with a transcriptional profile matching profibrotic monocyte-derived alveolar macrophages were observed in all three patients with pulmonary fibrosis, but not in the donor lungs (Figure 4d-f).

Profibrotic macrophages are thought to activate and stimulate the proliferation of fibroblasts via the generation of self-sustaining circuits maintained by the release of M-CSF from fibroblasts and factors promoting fibroblast activation and growth from macrophages^16,17^. In two recent single cell RNA-Seq atlases, a population of myofibroblasts was expanded in patients with pulmonary fibrosis compared with controls^12,13^. We observed a similar population marked by high levels of expression of *CTHRC1* and *POSTN* in the lungs from patients with COVID-19. Similarly, we observed that a population of fibroblasts expressing *HAS1* was present in COVID-19 patients^13^. We confirmed the presence of HAS1 positive fibroblasts colocalized with abnormal basaloid cells in the distal lung of explanted tissue (Figure 3g-i). Using immunofluorescence, we confirmed that KRT17 expressing epithelial cells are found in the lung periphery near fibroblasts (marked by COL1A1) in both explants (Figure 4j-l).

## DISCUSSION

We performed successful lung transplantation on two patients with respiratory failure secondary to SARS-CoV-2 pneumonia. Both patients had severe disease, requiring ECMO to maintain adequate oxygenation. Furthermore, both patients had shared complications of the SARS-CoV-2 infection complicating prolonged supportive ICU care. These included markedly reduced lung compliance, repeated episodes of ventilator associated pneumonia with increasingly resistant nosocomial pathogens, pneumothoraces requiring repeated tube thoracostomy, and bleeding into both the pleural space and airways. In addition, the pathology from the explants showed extensive and severe damage in both samples, again with significant similarities. Both patients showed diffuse alveolar damage, areas of bronchopneumonia with necrosis, extensive pleural inflammation, rare microthrombi, alveolar hemorrhage, and the accumulation of hemosiderin labeled alveolar macrophages and occasional giant cells in the lung. In addition, both patients showed extensive and bilateral cysts in the lung that were visible on microscopic sections and sometimes grossly. Microscopic examination revealed these cysts were extensive, variable in size and were lined by dysplastic cuboidal or sometimes squamous epithelium. In the surrounding interstitium, the normal alveolar architecture was largely lost with extensive interstitial thickening and fibrosis with areas suggestive of dense organizing pneumonia. The pathological findings suggest that the damage and destruction to the lungs were irreversible, and transplant was the sole viable treatment option for survival.

Given that the duration of SARS-CoV-2 infection in the lungs of patients with severe disease is uncertain, we were concerned about ongoing infection at the time of transplantation and re-infection of the allograft, particularly in our first patient who had received immunosuppression with rituximab and mycophenolate prior to her illness. Accordingly, we performed repeated bronchoscopic sampling of multiple lung regions prior to transplant and tested them using PCR. Reassuringly, we did not detect viral transcripts either by single cell RNA-Seq or by smFISH, suggesting bronchoscopic sampling is a clinically useful method to exclude SARS-CoV-2 infection prior to consideration for lung transplant. Our data are consistent with recent studies suggesting it is rare to detect replicating virus more than 10 days after infection with SARS-CoV-2 ^18-20^.

An emerging literature suggests a model for pulmonary fibrosis in which injury, often viral, to the alveolar epithelium triggers the recruitment monocyte-derived alveolar macrophages to the alveolar space^21^. These macrophages function to seal the wound, in the process forming reciprocal circuits with interstitial fibroblasts in which macrophage-produced factors stimulate fibroblast proliferation and matrix production, while fibroblast produced CSF1 maintains monocyte-derived alveolar macrophages in the niche^17^. Meanwhile, alveolar type 2 cells, a local alveolar epithelial progenitor cell, differentiate into type 1 cells in response to incompletely understood signals from the mesenchyme^22^. Failed repair allows macrophage-fibroblast circuits to persist, eventually filling the space with matrix proteins and fibroblasts. At the level of single cell RNA-Seq, we observed similarities between the end stages of pulmonary fibrosis and SARS-CoV-2 pneumonia. Specifically, we observed the emergence of an abnormal population of basaloid like epithelial cells that have been observed lining fibroblastic foci in patients with idiopathic pulmonary fibrosis (IPF), as well as profibrotic macrophages, myofibroblasts and *HAS1* expressing fibroblasts^9,12,13^. Furthermore, both patients with IPF and end stage SARS-CoV-2 pneumonia showed increased and disorganized matrix deposition. However, it is likely that the pathogenesis of pulmonary fibrosis in these patients is multifactorial and contributed by cytopathic effects of the virus, inflammation, barotrauma, and secondary bacterial pneumonias. Nevertheless, collectively these data suggest some patients develop an irreversible fibrotic lung disease for which transplantation is likely the only option for survival. The recovery observed in our patients also suggests that that in carefully selected individuals, lung transplantation can be utilized successfully despite the neuromuscular deconditioning that may result from COVID-19 acute respiratory distress syndrome.

## Data Availability

Counts tables and integrated objects are available through GEO with accession number GSE158127. Raw data is in the process of being deposited to SRA/dbGaP.

## ACKNOWLDEGMENTS

The authors have no conflict of interest to declare. This work was supported by the National Institutes of Health, NIH HL145478, HL147290, and HL147575 to AB; NIH U19AI135964, P01AG049665, R01HL153312, NUCATS COVID-19 Rapid Response Grant, CZI Seed Networks for the Human Cell Atlas to AVM, and P01AG049665, R01HL147575, U19AI135964, and Veterans Affairs grant I01CX001777 to GRSB. We thank pathology residents Taylor Zak, MD, PhD, and Megan Kinn, DO, for processing the human lungs for gross lung images.

Northwestern University Flow Cytometry Core Facility and Northwestern Center for Advanced Microscopy are supported by NCI Cancer Center Support Grant P30 CA060553 awarded to the Robert H. Lurie Comprehensive Cancer Center.

This research was supported in part through the computational resources and staff contributions provided by the Genomics Compute Cluster which is jointly supported by the Feinberg School of Medicine, the Center for Genetic Medicine, and Feinberg’s Department of Biochemistry and Molecular Genetics, the Office of the Provost, the Office for Research, and Northwestern Information Technology. The Genomics Compute Cluster is part of Quest, Northwestern University’s high-performance computing facility, with the purpose to advance research in genomics.

## METHODS

### Human Subjects

This study was approved by the Institutional Review Board of Northwestern University STU00212120, STU00213177, STU00212511 and STU00212579.

### Patients

#### Lung Transplant Patients

Described in the main text.

#### Post-mortem biopsy patient 1

A man in his 50s with medically controlled hyperlipidemia presented with five days of dyspnea, cough, and diarrhea. A nasopharyngeal swab was positive for SARS-CoV-2. He was intubated for ventilator management was guided by ARDSnet guidelines and the patient underwent three rounds of proning. However, he developed worsening hypoxemia and hypercapnia, necessitating initiation of veno-venous ECMO. Throughout the ECMO support, he underwent verticalization therapy to improve lung recruitment. His medical course spanning 29 days was complicated with acute renal failure, spontaneous hemothorax, pulmonary hemorrhage, and ventilator associated pneumonia. He tested negative for SARS-CoV-2 using the bronchoalveolar fluid on day 27^th^ from symptom onset for the first time. He received hydroxychroloquine, Remdesivir, and Sarilumab. However, he progressed to multiorgan dysfunction and care was withdrawn following which he expired within 40 minutes. Within 1 hour of death, warm autopsy of the left lung was performed. The tissue was processed for single cell RNAseq and SHIELDS tissue clearing.

#### Post-mortem biopsy patient 2

A woman in her 50s with no known medical conditions presented with dyspnea and was initially treated with one week course of steroids and hydroxychloroquine. She was intubated for severe ARDS and managed with prone ventilation based on ARDS net guidelines. Following intubation, she received Remdesivir but she continued to worsen requiring VV ECMO. She received broad-spectrum antibiotics and systemic anticoagulation throughout her course. She was tested negative for SARS-CoV-2 on day 31 in the bronchoalveolar fluid. However, the medical course was complicated by multiorgan multipressor shock. On day 48 after onset of symptoms, the medical care was redirected to comfort care and the patient expired within 10 minutes. Within 1 hour of expiration, left lung was biopsied and processed for single cell RNAseq and SHIELDS tissue clearance.

#### Autopsy specimen (smFISH for SARS-CoV-2)

An woman in 80s with end stage renal disease and cirrhosis was admitted for a fever of 101.3F. She had a positive nasopharyngeal swab for SARS-CoV-2. The patient developed increased O2 requirements and was subsequently transferred to the COVID ICU. In the ICU, the decedent developed hypotension to 60s/40s and after discussion with the clinical team, the decedent’s family elected to focus on comfort care. The patient died 8 days after admission.

#### ICU management

Lung transplantation Case 2 received most of his care at a referring hospital where the details of management are not known. At our institution, he and the other three patients were managed according to a standardized care plan for patients with COVID-19 that mirrors care for other patients with ARDS. Specifically, all patients received mechanical ventilation according to ARDSnet criteria (low VT, high PEEP)^1^. Pts received prone ventilation when the PaO2/FIO2 ratio was <150, repeated for 16 hours daily until criteria were no longer met^2^. Patients underwent bronchoalveolar lavage when pneumonia was suspected and antibiotic therapy was guided by quantitative cultures or the results of multiplex PCR analysis of the fluid (BioFire FilmArray Respiratory Viral Panel 2).

#### Lung Biopsy Tissue Fixation

Human lung biopsies collected during native lung explant or during post-mortem biopsy were sliced to 5 mm thickness and placed in 4% paraformaldehyde solution for 72 hours at room temperature. Samples were then exchanged to 70% ethanol and stored in 4°C until use. Biopsies were also sent to the clinical pathology laboratories where they were fixed, paraffin embedded and stained according to standard clinical protocols.

#### Lung biopsy tissue thick slicing

Biopsy samples were sliced to 100 µm thickness using a VT1200S Leica vibratome at 3.0 mm amplitude and 0.7 mm/s speed. For healthy lung biopsies, 100 µm thick slices were made at 3.0 mm amplitude and 0.35-0.4 mm/s speed with ice in the holding chambers. Slices were stored in a tissue culture plate in 1X phosphate buffered saline (PBS) with 0.2% sodium azide, the tops and sides were sealed in parafilm. Plates were stored in 4°C until use.

#### RNAscope of paraffin lung slices

RNAscope Multiplex V2 manual assay from ACDbio was performed on paraffin embedded 5 µm slices of lung tissue using mild digest times according to manufacturer instructions as we have described^3^. Probes used were RNAscope Probe-V-nCoV2019-S-C3 (catalog number 848561) with Akoya Bio Opal Dye 520 using the 488 laser line and RNAscope Probe-nCoV2019-orf1ab-sense-C2 (catalog number 859151) with Opal Dye 690 using 640 laser line. After RNAscope assay was complete, slides were washed in TBST (1X TBS pH 7.6 with 500 µL Tween-20) for 2 minutes with agitation twice. Slides were incubated in the dark at room temperature for 30 minutes with 10% normal goat serum in 1X TBS with 1% bovine serum albumin (BSA). Blocking solution was removed from slides via flicking. Slides were then incubated in primary antibody solution using CD206 Antibody (C-10) AF546 from Santa Cruz Biotechnology (RRID:AB_10989352) at 1:100 dilutions in TBS 1% BSA for 1h at room temperature in the dark. Slides were rinsed using TBST for 5 minutes with agitation twice. Slides were rinsed in TBST buffer for 5 minutes twice. Slides were then mounted and dried overnight. Images were taken in the Center for Advanced Microscopy at Northwestern University Feinberg School of Medicine using the Nikon W1-Spinning Disk Confocal microscope. Nucleus was added to the images using machine-based learning network trained on one patient using DAPI and brightfield images in Nikon Elements. Final images were rendered using Fiji.

#### SHIELD fixation of lung tissue slices

The 100 µm lung slices were secondarily fixed using a derivation of the SHIELD fixation protocol^4^. Slices were placed in clean 12-well tissue culture plate with 1 mL of SHIELD-Off solution (1:1:2 of ddH2O: SHIELD Buffer: SHIELD Epoxy). Plate was incubated at 4°C for 4.8 h with agitation. Slices were transferred to a clean plate containing 1 mL of SHIELD-On solution (1:1 SHIELD-On Buffer: SHIELD Epoxy) and incubated for 2.4 h at room temperature with agitation. Slices were cleared using passive methods, slices were placed in clean 12-well plate with LifeCanvas Passive Clearance Buffer at 37°C with agitation until slices were opaque; for healthy lung tissue incubate for 30-60 min, for diseased lung slices incubate for approximately 4 hours. Slices were washed overnight in PBS with 1% Triton-X-100 (PBST). Slices were stained in 1:10,000 dilution of Hoescht 33342 in PBST overnight. Slices were washed three times for 20 min in PBST after staining. Slices were placed in LifeCanvas Easy Index solution in a clean tissue culture plate for index matching.

#### Imaging of cleared lung tissue

The cleared tissue slices were imaged in the Center for Advanced Microscopy at Northwestern University Feinberg School of Medicine using the Nikon W1-Spinning Disk Confocal microscope in a glass bottom dish at 20X magnification. The 3D images were rendered using Fiji.

#### Single cell RNA-Seq of lung tissue

Single cell RNA sequencing was performed using modifications to our published protocols^3^. Distal lung biopsies were obtained from the explanted lung and donor lung from Case 1, and the two post-mortem biopsies. In addition, a biopsy that included the main left bronchus and distal parenchyma from the upper lobe was obtained from another donor lung that was not placed for transplant. Lung and airway tissues were infused with a solution of Collagenase IV and DNase I in RPMI, cut into ∼2 mm pieces and incubated in 10 ml of digestion buffer with mild agitation for 30 min. The Resulting single cell suspension was filtered through a 70 µm nylon mesh filter and digestion was stopped by addition of 10 ml of PBS supplemented with 0.5% BSA and 2 nM EDTA (staining buffer). Cells were pelleted by centrifugation at 300 rcf for 10 min, supernatant was removed and erythrocytes were lysed using 5 ml of 1x Pharm Lyse solution (BD Pharmingen) for 3 min. The single cell suspension was resuspended in Fc-Block (Human TruStain FcX, Biolegend) and incubated with CD31 microbeads (Miltenyi Biosciences; 130-091-935) and the positive fraction, containing endothelial cells and macrophages) was collected. The negative fraction was then resuspended in staining buffer, the volume was adjusted so the concentration of cells was always less than 5×10^7^ cells/ml and the fluorophore-conjugated antibody cocktail was added in 1:1 ratio (EpCAM, Clone 9C4, PECy7, BioLegend Cat# 324222, RRID:AB_2561506, 1:40; CD206, Clone 19.2, PE, ThermoFisher Cat# 12-2069-42, RRID:AB_10804655, 1:40; CD31, Clone WM59, APC, BioLegend Cat# 303116, RRID: AB_1877151, 1:40; CD45 Clone HI30, APCCy7, BioLegend Cat# 304014, RRID: AB_314402, 1:40; HLA-DR, Clone LN3, eFluor450, ThermoFisher Cat# 48-9956-42, RRID:AB_10718248, 1:40). After incubation at 4C for 30 min cells were washed with 5 ml of MACS buffer, pelleted by centrifugation and resuspended in 500 ul of MACS buffer + 2 ul of SYTOX Green viability dye (ThermoFisher). Cells were sorted on FACS Aria II SORP instrument using a 100 um nozzle and 20 psi pressure. Macrophages were sorted as live/CD45^+^HLA-DR^+^CD206^+^ cells, epithelial cells were sorted as live/CD45^−^CD31^−^EpCAM^+^, stromal cells were sorted as live/CD45^−^CD31^−^EpCAM^−^ cells. Sample processing was performed in BSL-2 conditions using BSL-3 practices. Cells were sorted into 2% BSA in DPBS, pelleted by centrifugation at 300 rcf for 5 min at 4C, resuspended in 0.1% BSA in DPBS to ∼1000 cells/ul concentration. Concentration was confirmed using K2 Cellometer (Nexcelom) with AO/PI reagent and ∼5,000–10,000 cells were loaded on a 10x Genomics Chip B with Chromium Single Cell 3’ gel beads and reagents (3’ GEX V3, 10x Genomics). Libraries were prepared according to the manufacturer’s protocol (10x Genomics, CG000183_RevB). After quality check single cell RNA-seq libraries were pooled and sequenced on a HiSeq4000 or NovaSeq 6000 instrument. Data was processed using Cell Ranger 3.1.0 pipeline (10x Genomics). To enable detection of viral RNA, reads were aligned to a custom hybrid genome containing GRCh38.93 and SARS-CoV-2 (NC_045512.2). Data was processed using Scanpy v1.5., doublets were detected with scrublet v0.2.1 and removed, ribosomal genes were removed and multisample integration was performed with BBKNN v1.3.12. Detailed analysis code is available at https://github.com/NUPulmonary/2020_Bharat. Counts tables and integrated objects are available through GEO with accession number GSE158127. Raw data is in the process of being deposited to SRA/dbGaP.

## REFERENCES

1. Geleris J, Sun Y, Platt J, et al. Observational Study of Hydroxychloroquine in Hospitalized Patients with Covid-19. N Engl J Med 2020;382:2411–8.

2. Richardson S, Hirsch JS, Narasimhan M, et al. Presenting Characteristics, Comorbidities, and Outcomes Among 5700 Patients Hospitalized With COVID-19 in the New York City Area. JAMA 2020.

3. Group RC, Horby P, Lim WS, et al. Dexamethasone in Hospitalized Patients with Covid-19 - Preliminary Report. N Engl J Med 2020.

4. Beigel JH, Tomashek KM, Dodd LE, et al. Remdesivir for the Treatment of Covid-19 - Preliminary Report. N Engl J Med 2020.

5. van der Mark SC, Hoek RAS, Hellemons ME. Developments in lung transplantation over the past decade. Eur Respir Rev 2020;29.

6. Brower RG, Lanken PN, MacIntyre N, et al. Higher versus lower positive end-expiratory pressures in patients with the acute respiratory distress syndrome. N Engl J Med 2004;351:327–36.

7. Guérin C, Reignier J, Richard J-C, et al. Prone Positioning in Severe Acute Respiratory Distress Syndrome. New England Journal of Medicine 2013;368:2159–68.

8. Ackermann M, Verleden SE, Kuehnel M, et al. Pulmonary Vascular Endothelialitis, Thrombosis, and Angiogenesis in Covid-19. N Engl J Med 2020;383:120–8.

9. Reyfman PA, Walter JM, Joshi N, et al. Single-Cell Transcriptomic Analysis of Human Lung Provides Insights into the Pathobiology of Pulmonary Fibrosis. Am J Respir Crit Care Med 2019;199:1517–36.

10. Vaughan AE, Brumwell AN, Xi Y, et al. Lineage-negative progenitors mobilize to regenerate lung epithelium after major injury. Nature 2015;517:621–5.

11. Habiel DM, Espindola MS, Jones IC, Coelho AL, Stripp B, Hogaboam CM. CCR10+ epithelial cells from idiopathic pulmonary fibrosis lungs drive remodeling. JCI Insight 2018;3.

12. Adams TS, Schupp JC, Poli S, et al. Single-cell RNA-seq reveals ectopic and aberrant lung-resident cell populations in idiopathic pulmonary fibrosis. Science advances 2020;6:eaba1983.

13. Habermann AC, Gutierrez AJ, Bui LT, et al. Single-cell RNA sequencing reveals profibrotic roles of distinct epithelial and mesenchymal lineages in pulmonary fibrosis. Science advances 2020;6:eaba1972.

14. Misharin AV, Morales-Nebreda L, Reyfman PA, et al. Monocyte-derived alveolar macrophages drive lung fibrosis and persist in the lung over the life span. The Journal of Experimental Medicine 2017.

15. Valenzi E, Bulik M, Tabib T, et al. Single-cell analysis reveals fibroblast heterogeneity and myofibroblasts in systemic sclerosis-associated interstitial lung disease. Annals of the Rheumatic Diseases 2019;78:1379–87.

16. Joshi N, Watanabe S, Verma R, et al. A spatially restricted fibrotic niche in pulmonary fibrosis is sustained by M-CSF/M-CSFR signalling in monocyte-derived alveolar macrophages. Eur Respir J 2020;55.

17. Zhou X, Franklin RA, Adler M, et al. Circuit Design Features of a Stable Two-Cell System. Cell 2018;172:744-57.e17.

18. Wolfel R, Corman VM, Guggemos W, et al. Virological assessment of hospitalized patients with COVID-2019. Nature 2020;581:465–9.

19. Arons MM, Hatfield KM, Reddy SC, et al. Presymptomatic SARS-CoV-2 Infections and Transmission in a Skilled Nursing Facility. N Engl J Med 2020;382:2081–90.

20. Bullard J, Dust K, Funk D, et al. Predicting infectious SARS-CoV-2 from diagnostic samples. Clin Infect Dis 2020.

21. Misharin AV, Morales-Nebreda L, Reyfman PA, et al. Monocyte-derived alveolar macrophages drive lung fibrosis and persist in the lung over the life span. J Exp Med 2017;214:2387–404.

22. Juul NH, Stockman CA, Desai TJ. Niche Cells and Signals that Regulate Lung Alveolar Stem Cells In Vivo. Cold Spring Harbor perspectives in biology 2020.

